# Standard and anomalous waves of COVID-19: A multiple-wave growth model for epidemics

**DOI:** 10.1101/2021.01.31.21250867

**Authors:** Giovani L. Vasconcelos, Arthur A. Brum, Francisco A. G. Almeida, Antônio M. S. Macêdo, Gerson C. Duarte-Filho, Raydonal Ospina

**Affiliations:** Departamento de Física, Universidade Federal do Paraná, 81531-990 Curitiba, Paraná, Brazil; Departamento de Física, Universidade Federal de Pernambuco, 50670-901 Recife, Pernambuco, Brazil; Departamento de Física, Universidade Federal de Sergipe, 49100-000, São Cristóvão, Sergipe, Brazil; Departamento de Estatística, Universidade Federal de Pernambuco, 50740–540, Recife, Pernambuco, Brazil

## Abstract

We apply a generalized logistic growth model, with time dependent parameters, to describe the fatality curves of the COVID-19 disease for several countries that exhibit multiple waves of infections. In the case of two waves only, the model parameters vary as a function of time according to a logistic function, whose two extreme values, i.e., for early and late times, characterize the first and second waves, respectively. For the multiple-wave model, the time-dependency of the parameters is likewise described by a multi-step logistic function with *N* intermediate plateaus, representing the *N* waves of the epidemic. We show that the theoretical curves are in excellent agreement with the empirical data for all countries considered here, namely: Brazil, Canada, Germany, Iran, Italy, Japan, Mexico, South Africa, Sweden, and US. The model also allows for predictions about the time of occurrence and severity of the subsequent waves. It is shown furthermore that the subsequent waves of COVID-19 can be generically classified into two main types, namely, standard and anomalous waves, according as to whether a given wave starts well after or well before the preceding one has subsided, respectively.

## 1 Introduction

More than one year after the first death by the novel coronavirus (SARS-CoV-2), on January 11, 2020, in Wuhan, China, the direst predictions about the danger and severity of the ensuing pandemic have been confirmed. As of this writing, more than 143 millions of cases of infection by the SARS-CoV-2 virus haven been identified worldwide and over 3 millions of deaths have been attributed to the disease (COVID-19) caused by the virus^1, 2^. The response strategies to counter the propagation of the virus have varied widely from country to country, and even within countries. Notwithstanding their different approaches to fight the COVID-19 pandemic, a great deal of countries have suffered a significant loss of life to the virus. A particularly interesting but troublesome development in the course of the pandemic is the fact that many countries were able to control somewhat the spread of the disease during the first few months after its onset, only to see a subsequent increase in the rate of infections and deaths. This resurgence of the COVID-19 epidemic, commonly referred to as a second wave of infections, has in some cases been even more severe that the so-called first wave. Furthermore, in several countries successive waves have been observed beyond the second resurgence of the epidemic—in some cases up to a fourth wave of infections. Hence describing the multiple-wave dynamics of the COVID-19 pandemic is a topic of special relevance, both from the mathematical modeling viewpoint and from the public health perspective.

In the context of the COVID-19 epidemic, a second wave of infections can, broadly speaking, appear via two main different dynamics. First, in a standard or textbook second wave, the resurgence of infections appears after the epidemic curves for the cumulative number of cases and deaths have reached a near-plateau. This means that the daily numbers of new infections and deaths have decreased substantially—and remains low for a somewhat prolonged period of time—, before they surge again. Correspondingly, the daily curves display two well-defined sharp “peaks,” separated by a rather shallow “valley.” In other words, in a standard second wave there is a clear, distinct separation between the first and the second waves of infections. There are other situations, however, where the epidemic curve undergoes a strong re-acceleration regime even before the daily rates of infections and death have been significantly reduced, indicating that a second wave starts before the first wave has subdued. Such an “anomalous” second-wave effect shows up in the respective cumulative curve as a rapid change in the trend of the growth profile, i.e., from deceleration to acceleration, even though no plateau-like regime had yet been reached. Thus, in an anomalous second wave there is a transition period where the first and the second waves can be said to “coexist,” as represented by the fact that the two peaks in the daily curve are separated by a relatively high valley. A similar qualitative classification can be applied to third and subsequent waves.

Locating and quantifying multiple-wave effects in a given epidemic curve, beyond simple visual inspection, is an important but not a trivial task. This requires, for instance, a mathematical or computational model that is able to efficiently describe the complex growth profiles that arise in the cumulative epidemic curves and from which one can estimate the location and intensity of each wave’s peak in the daily curves. A standard way to investigate multiple-wave effects is to start with a basic epidemiological model and then allow its parameters to vary in time to reflect the occurrence of secondary waves of infections^3, 4^. Understanding the evolution of possible multiple waves of infections is also, of course, relevant for public health officials, as it may help them to develop better strategies to fight the propagation of the virus. In response to the widespread occurrence of second and subsequent waves of the COVID-19 epidemic in many countries around the world, there is now a fast growing body of literature on the subject^5–22^. In such studies, compartmental models^9–11^ and agent based models^14, 15^ are typically the models of choice, although models based on artificial intelligence algorithms have also been used^16–19^, as well as observational data^20–22^.

In this paper, we depart from previous approaches and propose to model multiple-wave effects in the COVID-19 epidemic in terms of a generalized logistic model with time-dependent parameters. More specifically, we consider an extension of the so-called beta logistic model (BLM)^23^, where we assume that each parameter of the model (see below) is allowed to vary continuously and smoothly in time between a number *N* of well defined values, representing the *N* waves of the epidemic dynamics. We apply the model to study the fatality curves of COVID-19, as represented by the cumulative number of deaths as a function of time, for ten selected countries that display second and third waves, namely: Brazil, Canada, Germany, Iran, Italy, Japan, Mexico, South Africa, Sweden, and US. We show that the generalized BLM describes very well the mortality curves of all selected countries. Furthermore, from the theoretical curves we are able to quantify the transition times between the successive waves as well as the relative intensity of a subsequent wave relative to the preceding one.

## 2 Data

Here we focus exclusively on mortality data from COVID-19, rather than on the number of infection cases. The reason for this choice is because it is difficult to estimate the actual number of infected people by the SARS-CoV-2, since the confirmed cases represent only an unknown fraction of the total number of infections. In this scenario, the number of deaths attributed to COVID-19 is a somewhat more reliable measure to describe the dynamics of the epidemic^24^.

As our main aim here is to analyze the second and third waves of the COVID-19 epidemic in different countries, we have included only data from countries that, up to the maximum date considered here, namely April 03, 2021, displayed at least two and at most three waves of infection. More specifically, we have analyzed the COVID-19 fatality curves for ten selected countries, namely: Brazil, Canada, Germany, Iran, Italy, Japan, Mexico, South Africa, Sweden, and US.

The data used in this study were obtained from the database made publicly available by the Johns Hopkins University^1, 25^, which lists in automated fashion the number of the confirmed cases and deaths per country. As already mentioned, in all cases considered here we have considered COVID-19 mortality data up to April 03, 2021.

## 3 Methods

In the present paper we are mainly interested in modeling epidemic curves that display multiple-wave effects, as indicated for example by the presence of two or more regimes with strong positive accelerations, corresponding to the first and subsequent waves of infection. Because our model for such cases is a generalization of a single-wave model, we shall start by reviewing the basic one-wave model (i.e., with constant parameters), after which the general model with time-dependent parameters is discussed. Subsequently, we also discuss the numerical methods used to analyze the empirical data.

### 3.1 Single-Wave Model

We model the time evolution of the number of deaths in the epidemic by means of the beta logistic model (BLM), defined by the following ordinary differential equation (ODE)^23, 26^:

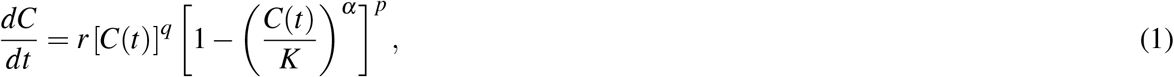

where *C*(*t*) is the cumulative number of deaths at time *t*. We assume for the time being that the model parameters {*r, q, α, p, K*} are all constant in time, in which case they can be interpreted as follows: *r* is the growth rate at the early stage; *q* controls the initial growth profile and allows to interpolate from linear growth (*q* = 0) to sub-exponential growth (*q* < 1) to purely exponential growth (*q* = 1); the exponent *p* controls the late-time growth rate, with *p* > 1 implying a slow-decaying polynomial rate, whereas *p* = 1 yields a fast exponential decay (see below); the exponent *α* controls the degree of asymmetry with respect to the symmetric S-shape of the logistic curve, which is recovered for *q* = *p* = *α* = 1; and, finally, *K* is the final size of the epidemic, meaning that *C*(*t*) = *K*, for *t* → ∞. Equation (1) must be supplemented with the initial condition

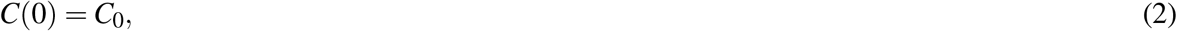

for some given value of *C*_0_.

The BLM described in (1) is one of the most general growth models, from which many other known models emerge as special cases^23, 26^. For instance, for *q* = *p* = *α* = 1 we recover the standard logistic model, as already mentioned. In addition, for *q* = *p* = 1 we obtain the Richards growth model^27^, with the case *p* = 1 corresponding to the so-called generalized Richards model^28^; while setting *α* = 1 in (1) yields the Blumberg’s equation^29^; for other special cases see Ref.^26^.

In the case where the parameters {*r, q, α, p, K*} are constant, the BLM admits an analytic solution^23^ in the following implicit form:

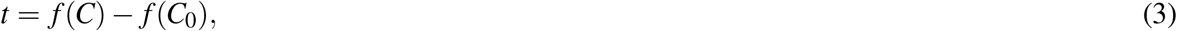

where

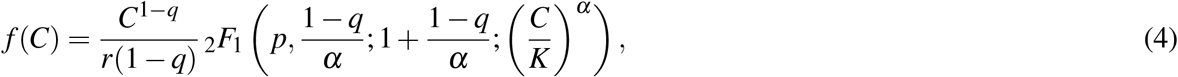

with _2_*F*_1_(*a, b*; *c*; *x*) being the Gauss hypergeometric function. Equation (3) describes a sigmoidal curve, whose only inflection point is located at the time *t*_*c*_ obtained by substituting the value *C*_*c*_ = *K*[*q/*(*q* + *αp*)]^1*/α*^ in (3). The small- and large-times asymptotic behavior of the growth profile *C*(*t*) are as follows^23^:

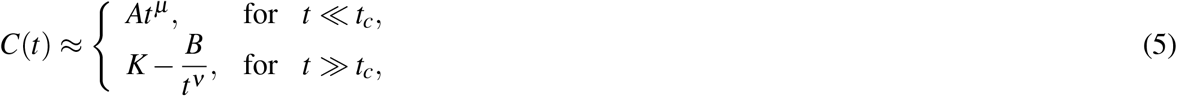

where *µ* = 1*/*(1 −*q*), *A* = [*r*(1 −*q*)]^1*/*(1−*q*)^, *ν* = 1*/*(*p* − 1), and *B* = [*K*^*p*−*q*^*/*(*p*− 1)*rα*^*p*^] ^1*/*(*p*−1)^. (For *q* → 1 and *p* → 1, one obtains exponential growth and exponential decay, respectively.)

Growth models have the mathematical advantage that they often admit analytical solutions, as we have shown above for the BLM, which is a very useful property when fitting models to empirical data^30^. Furthermore, it is worth pointing out that there is an intrinsic connection between growth models and mechanistic epidemic models of the Susceptible-Infected-Recovered (SIR) class of models. For instance, it is possible to construct a map between the Richards growth model and SIR-type models^31, 32^. Thus, when properly applied and interpreted, growth models are useful tools for understanding the spreading dynamics of infectious diseases^23, 24, 28^. Although in the present paper we restrict our analysis to the BLM, it is important to bear in mind the aforementioned connection between growth models and compartmental models. We therefore thought it was worth including here a brief discussion about a map between the BLM and a generalized SIRD model.

### 3.2 SIRD Model with Power-Law Behavior

As mentioned above, it is possible to put the BLM in correspondence with a SIRD-like model, but in this case the target SIRD-type model has to be modified by the inclusion of a power law in the incidence term, owing to the power-law behavior exhibited by the BLM, as shown below.

We start by recalling the standard Susceptible (*S*)-Infected (*I*)-Recovered (*R*)-Deceased (*D*) epidemiological model^33, 34^

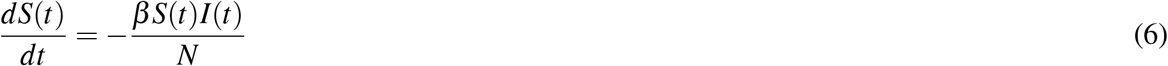

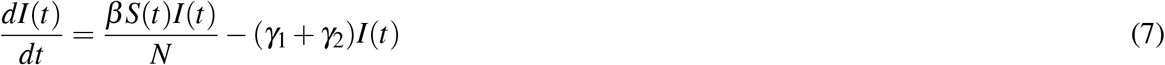

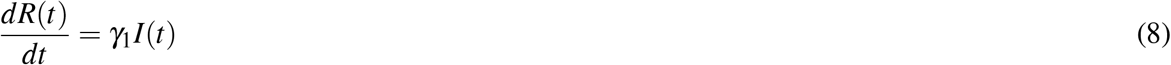

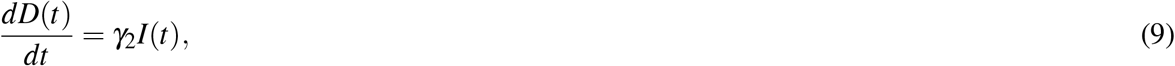

where *S*(*t*), *I*(*t*), *R*(*t*), and *D*(*t*) are the number of individuals at time *t* in the classes of susceptible, infected, recovered, and dead respectively, while *N* is the constant total number of individuals in the population, so that *N* = *S*(*t*) + *I*(*t*) + *R*(*t*) + *D*(*t*). The parameters *β, γ*_1_ and *γ*_2_ are the transmission, recovery and death rates respectively. The initial values can be chosen to be *S*(0) = *S*_0_, *I*(0) = *I*_0_, with *S*_0_ + *I*_0_ = *N*, and *R*(0) = 0 = *D*(0).

We then consider a modified SIRD model, where in (6) and (7) we replace *N* with only the partial population in the *S* and *I* compartments^31^. Furthermore, we follow Refs.^35, 36^ and replace the term *I*(*t*) on the right-hand side of all equations above by [*I*(*t*)]^*p*^, to obtain

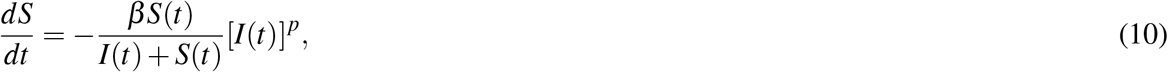

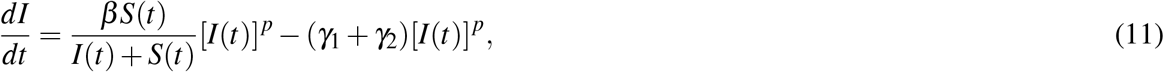

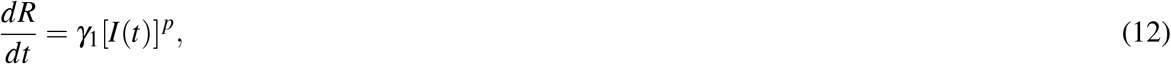

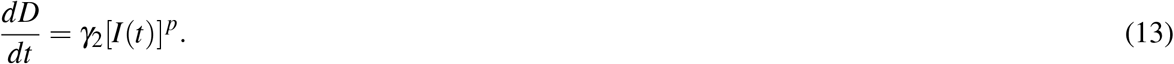

Although this model is still not general enough to accommodate all the phenomenology of the intervention biased dynamics of the COVID-19 epidemics, it does nonetheless exhibit subexponential behavior for both short and large time scales in all compartments. In order to show this, we define *y*(*t*) = *S*(*t*) + *I*(*t*) and divide (11) by (10) to obtain

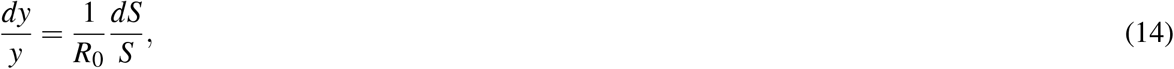

where *R*_0_ = *β/*(*γ*_1_ + *γ*_2_). Integrating both sides of (14), and inserting the result into (10), yields an equation of the BLM type:

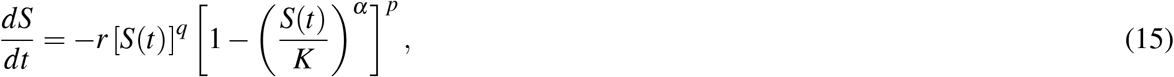

where *r, K* > 0, *q* = 1 + (*p* − 1)*/R*_0_ and *α* = 1 − 1*/R*_0_. It follows from (15), in comparison with model (1), that *S*(*t*) exhibits power-law regimes (for both early and large times) akin to those described by the BLM. Furthermore, it is easy to see that all other compartments, *I*(*t*), *R*(*t*), and *D*(*t*), inherit from *S*(*t*) the power-law behavior, even though their respective equations of motion are not of the BLM-type. Note that the parameters *q, p*, and *α* in (15) are not all independent of one another—in fact, the conditions *q* < 1 and *p* > 1 are mutually incompatible. Nonetheless, the preceding qualitative argument shows that the power-law dynamics of the sort predicted by the BLM can in principle be accommodated by compartmental models. We are currently carrying out further research to establish a more complete map between the BLM and a generalized SIRD model where the exponents *q, p*, and *α* are all independent of each other.

The BLM with constant parameters has been shown to describe remarkably well the first wave of the COVID-19 epidemic for several countries in Europe and North America^23^. However, after the resurgence of the COVID-19 epidemic in many countries (most notably after the Northern Hemisphere summer of 2020), their respective epidemic curves started to exhibit more complex patterns that cannot be captured by the standard BLM. In the next subsection we introduce a generalized version of the BLM with time-dependent parameters, which is much more adequate to describe growth processes with two or more distinct growth phases, corresponding to different waves of infection.

### 3.3 Multiple-Wave Model

Our multiple-wave model is still described by the ODE given in (1), but now we assume that *all parameters depend on time*, that is, *r* = *r*(*t*), *q* = *q*(*t*), *α* = *α*(*t*), *p* = *p*(*t*), and *K* = *K*(*t*). Let us first consider the case where there are only two waves of infections. To capture the two distinct growth regimes (corresponding to the first and second waves, respectively), we propose that these parameters, here generically represented by the symbol *ζ* (*t*), obey the following logistic-like equation

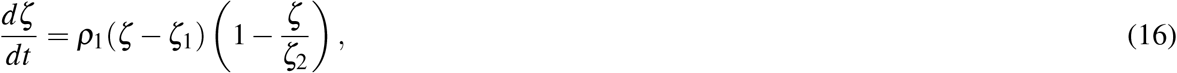

whose solution, with the condition *ζ* (*t*_1_) = (*ζ*_1_ + *ζ*_2_)*/*2, is of the following form:

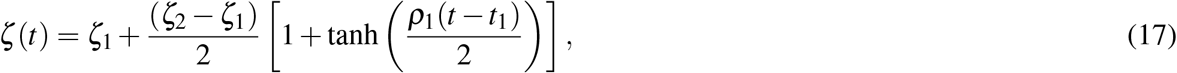

where *ζ* (*t*) stands for any of the parameters *r, α, q, p*, and *K*, with *ζ*_1_ and *ζ*_2_ representing the corresponding parameter values for the first and second waves, respectively. A schematic of the generic parameter *ζ* (*t*), as defined in (17), is shown in Fig. 1. The parameter *t*_1_ determines the transition time between the first and second wave; whereas the parameter *ρ*_1_ characterises how rapid this transition is, so that the larger the parameter *ρ*_1_ the quicker the transition towards the second-wave regime. Note that the characteristic time scale *t*_1_ and the corresponding transition rate *ρ*_1_ are the same for all parameters. This is justified because an overall change in the epidemic dynamics, brought about, say, by a relaxation of control measures or by a change in the population behavior (or both), is expected to affect simultaneously all epidemiological parameters, which are in turn described in an effective manner by the growth model parameters^24^.

**Figure 1.**
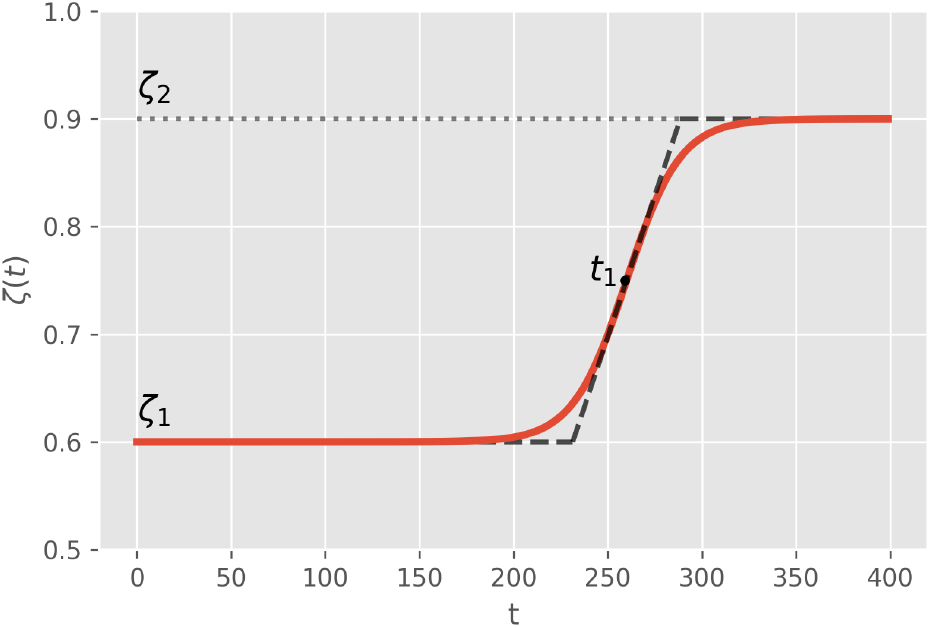
Time dependence of the generic parameter *ζ* (*t*) of the two-wave model, as defined by the logistic function given in (17). The dashed line represents the linear approximation to the logistic function, where the inclined straight line meets the upper and lower horizontal lines at the points *t*_1_ *±* 2*/ρ*_1_, where *t*_1_ and *ρ*_1_ are respectively the transition time and rate between the two plateaus; see text.

It is worth pointing out, however, that although the parameter *t*_1_ in (17) sets the time scale for the transition between the first and second waves, it does not, in itself, represent the time where the second-wave effects begin, since the onset of the second wave also depends on the parameter *ρ*_1_. This can been seen more clearly in Fig. 1, where we also indicated a piecewise linear approximation (dashed line) to the function (solid line) described by (17). Under this linear approximation, one sees that the linear transition region, *t*_1_ − 2*/ρ*_1_ < *t* < *t*_1_ + 2*/ρ*_1_, could be seen as a period when new fatalities can, at least from a theoretical viewpoint, be attributed both to the terminal phase of the first wave and the initial phase of the second wave. This superposition effect between successive waves could be avoided by imposing a discontinuous change of parameters, i.e., taking *ρ*_1_ → ∞, but in this case additional continuity condition on the derivative of *C*(*t*) is necessary, as considered, e.g., in Ref.^24^. For numerical purposes, it is more convenient however to describe the transition between the first and second waves with a smooth function, as indicated in (17). Physically, we believe that this smooth transition between different waves of infection is also more reasonable, as a resurgence of infections do not tend to occur suddenly. We anticipate, however, that estimates of the transition time computed from the linear approximation shown in Fig. 1, such as the lowermost point *t*_1_ − 2*/ρ*_1_, can result imprecise (for instance, if *r* is too small, this formula can largely underestimates the onset of the second wave). A better estimate for the onset time of the second wave will be presented below.

As an analytical solution for the theoretical curve *C*(*t*) for the BLM time-dependent parameters is no longer possible, one must resort to a numerical integration of the ODE (1), with the parameters {*r*(*t*), *q*(*t*), *α*(*t*), *p*(*t*), *K*(*t*)} described by their respective transition functions of the form given in (17). A schematic of the cumulative curve, *C*(*t*), for the two-model upon numerical integration (for an arbitrary choice of parameters) is shown in Fig. 2a. In this figure, the dashed line denoted by *K*_1_ represents the plateau level if only the first wave had been present, whereas the parameter *K*_2_ is the actual final plateau corresponding to the total number of deaths at the end of the epidemic (assuming that subsequent waves of infection do not occur). In Fig. 2b, we show the time derivative, *dC/dt*, of the cumulative curve shown in Fig. 2a, which corresponds to the daily number of deaths as a function of time. In Fig. 2b, the “peaks” of the two waves are indicated by the inverted triangles, with the respective values denoted by *P*_1_ and *P*_2_. Similarly, the minimum (“valley”) between the two peaks of the daily curve is denoted by *V*_1_ (black dot). This point is also indicated in Fig. 2a by a black dot, which marks the transition between the first and second waves in the cumulative curve. From figure 2, it is clear that it is convenient to take the time of occurrence of the minimum *V*_1_ as a practical criterion to estimate the beginning of the second wave.

**Figure 2.**
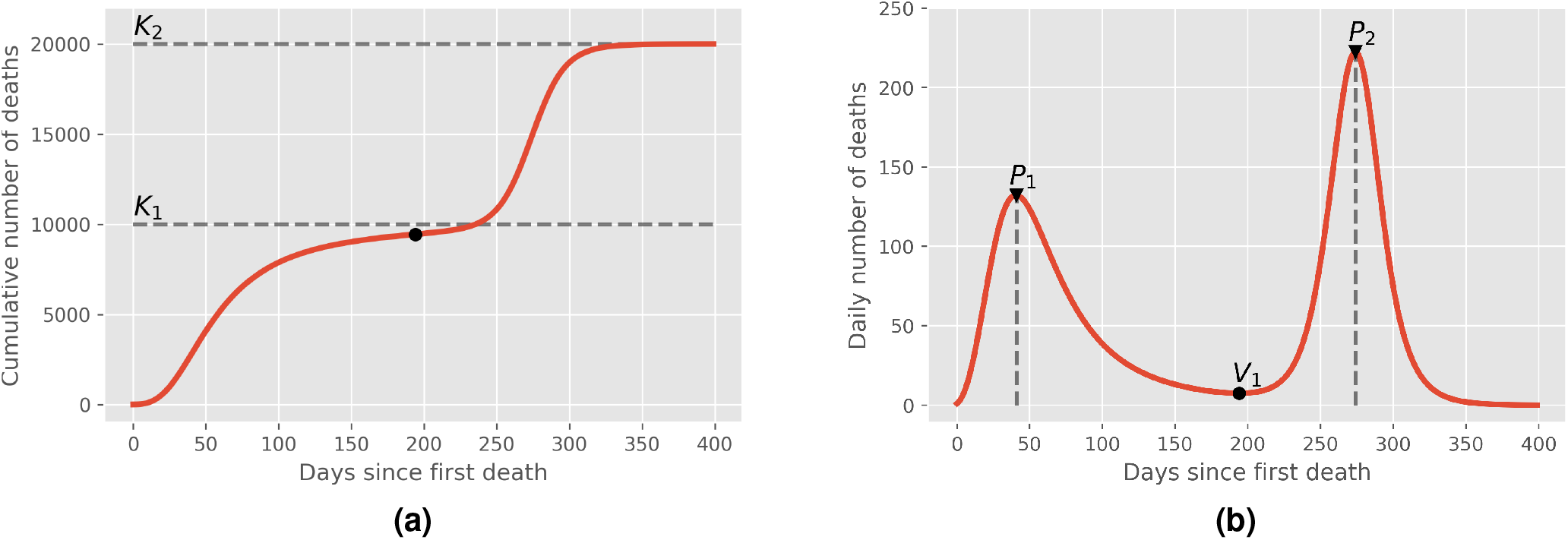
(a) Schematic of an epidemic curve for the cumulative number, *C*(*t*), of deaths with two waves of infections. Here *K*_1_ is the plateau value if only the first wave had been present and *K*_2_ is the actual value at the end of the epidemic (final plateau), assuming that no subsequent recrudescence of the epidemic occurs. (b) Time derivative, *dC/dt*, of the cumulative curve shown in (a), representing the daily number of deaths, where the peaks for the first and second waves are indicated by *P*_1_ and *P*_2_ (inverted triangles), respectively, and the minimum between these two peaks corresponds to *V*_1_ (black dot).

The two-wave model described above can be naturally extended to include subsequent multiple waves by considering time-dependent parameters of the following form:

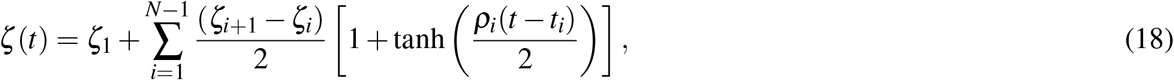

where *N* indicates the total number of infection waves within the epidemic. At face value, for a given number *N* of waves, our multiple-wave model has 5*N* + 2(*N* − 1) = 7*N* − 2 free parameters, corresponding to the initial and final values for each of the five BLM parameters {*r, q, α, p, K*}, together with the *N* − 1 parameters *t*_*i*_ and *ρ*_*i*_, *i* = 1, …, *N* − 1, describing the transition between successive waves. Blindingly trying to fit a given empirical epidemic curve with a model containing such a large number of parameters, even for the case of only two waves (for which there are 12 parameters to be determined), is not an efficient procedure, as one is bound to incur into over-fitting issues. Next, we describe a multiple-step fitting procedure that aims at circumventing, at least partially, these difficulties.

### 3.4 Data Analysis

In the first step of our fitting procedure, we give an initial educated guess for the possible location of the first transition time, *t*_1_, between the first and second waves. We then fit the data up to this time with the one-wave model, as given by its analytical solution (3). At this point, it is important to recall that the parameters *r, q*, and *α* in the one-wave model (1) are restricted to certain allowed ranges. For example, the exponent *q* is limited to the range 0 ≤ *q* ≤ 1, as *q* > 1 would imply a super-exponential growth which is not justified on epidemiological grounds. Furthermore, it is expected by biological reasons (see, e.g., the discussion in Refs.^24, 31^) that the asymmetry parameter *α* should also be within the interval (0,1). Similarly, we restrict the values of *r* to the range (0,1), as we observed that values of *r* outside this interval tends to be an indication of possible over-fitting. In other words, we assume here that the restrictions 0 < *q* ≤ 1, 0 < *α* ≤ 1, and 0 < *r* < 1 are useful empirical criteria to reduce over-fitting.

Next, we repeat the previous step up to the second wave, as follows. If the empirical curve in question has only two waves, we carry out the numerical fit of the entire data using the two-wave model, that is, eq. (18) with *N* = 2. If, however, the epidemic curve displays evidence of third-wave effects, we select an initial guess for the second transition time, *t*_2_, between the second and third waves, so that the fit with the two-wave model (*N* = 2) is applied only up to the time *t*_2_. In both cases, the values obtained for the parameters *r*_1_, *q*_1_, *α*_1_, *p*_1_, and *K*_1_ in the first step described above are used as initial guesses for the respective parameters describing the first wave in the two-wave model. The initial guesses for the second set of parameters, *r*_2_, *q*_2_, *α*_2_, *p*_2_, and *K*_2_, characterising the second wave, as well as for the rate of transition *ρ*_1_ are chosen somewhat arbitrarily within their ranges of definition. If the epidemic curve in question has only two waves, this second step concludes the numerical fitting. If, however, there is a tertiary wave in the data, we repeat the procedure above with the three-wave model (*N* = 3), whereby the two sets of parameters {*r*_*i*_, *q*_*i*_, *α*_*i*_, *p*_*i*_, *K*_*i*_}, for *i* = 1, 2, obtained in the previous step are used as the initial guesses for the corresponding parameters of full three-wave model, which is applied to the entire empirical data. As before, the initial guesses for the last set of parameters, namely, {*r*_3_, *q*_3_, *α*_3_, *p*_3_, *K*_3_}, are arbitrarily chosen in their range of validity.

This multiple-step procedure can in principle be applied to any number of epidemic waves. However, for large *N*, the numerical task of fitting a *N*-wave model (with 7*N* − 2 parameters) to a given empirical curve becomes quite challenging, specially considering that the total number of points in an empirical epidemic curve is relatively small (typically of the order of a few hundreds). Hence, in the present study we shall restrict ourselves to epidemic curves that have at most three waves.

In all numerical fits, for both the single-wave and multiple-wave models, we employed the Levenberg-Marquardt algorithm to solve the non-linear least square optimisation problem, as implemented in the `lmfit` package for the `Python` language, which has a built-in routine for estimating the errors of the fitted parameters via the covariance matrix^37^. The results of the fitting procedure are deemed acceptable when the errors in the parameters were smaller than the values themselves estimated for the parameters. In most cases reported here, however, the errors are much smaller than the maximum allowed tolerance of 100%. There were only two cases, namely South Africa and Italy, where the relative errors in some of the parameters for the corresponding last waves exceeded 100%. Nevertheless, we decided to keep these two examples here because the respective fits are still of good quality and also because estimating the model parameters associated with the last wave is inherently more difficult. This difficulty is a reflection of both over-fitting and the fact that there are generally fewer points in the final portions of the empirical curves, so that it is not surprising that the errors estimated for the parameters associated with the last wave tend to be larger. We noticed, furthermore, that in order to control the errors and minimize over-fitting issues, it was necessary to apply the restrictions *α*_2_ = 1 and *α*_3_ = 1 (in cases where there are three waves). This is because the asymmetry parameter *α*_*i*_ controls the bending towards the *i*-th plateau^23^, so that minor variations in these parameters may lead to large unacceptable errors in other parameters. Hence it proved convenient to set *α*_2_ = *α*_3_ = 1; but otherwise all other parameters are allowed to vary freely in their respective ranges of validity during the optimization procedure.

## 4 Results

As our main aim in this paper is to illustrate the application of our multiple-wave model, we have chosen a representative sample of fatality curves that have at least two and at most three regions of accelerated growth, which can be unmistakably associated with second and third waves of COVID-19 infections. With these goals in mind, we have analysed the COVID-19 fatality curves for ten selected countries, namely: Brazil, Canada, Germany, Iran, Italy, Japan, Mexico, South Africa, Sweden, and US. For all selected countries, we have included data up to April 03, 2021. Up to this date, six countries (among the selected ones) display two epidemic waves, namely: Brazil, Canada, Germany, Mexico, South Africa, and Sweden; while the other four countries, namely Iran, Italy, Japan, and US, already have three waves of infections. Below we shall present our results for each of these two sets of countries (i.e., with only two and with three waves) separately.

### 4.1 Countries with Two Waves

As already discussed in the Introduction, a second wave of infections can, broadly speaking, take place in two main different ways. First, a “standard” second wave pattern can be said to occur when the epidemic curve re-embarks on a rapid acceleration regime after the first wave of infections had nearly ‘died out.’ This means that the cumulative curve has reached a near-plateau, before it surges up again.

Among the six countries with two waves, three of them (Canada, Germany, and Sweden) displayed standard second waves, as shown in Fig. 3. In the left panels of this figure, we show the cumulative number of deaths (red circles) attributed to COVID-19 for these three countries, as a function of time counted in days since the first death in each country. Also shown in these figures are the corresponding best fits (black solid curves) by the two-wave model given in (1) and (17). One sees from the plots in Fig. 3 that the theoretical curves describe remarkably well the empirical data for all cases. The respective best-fit parameters are shown in the legend box of each graph in Fig. 3.

**Figure 3.**
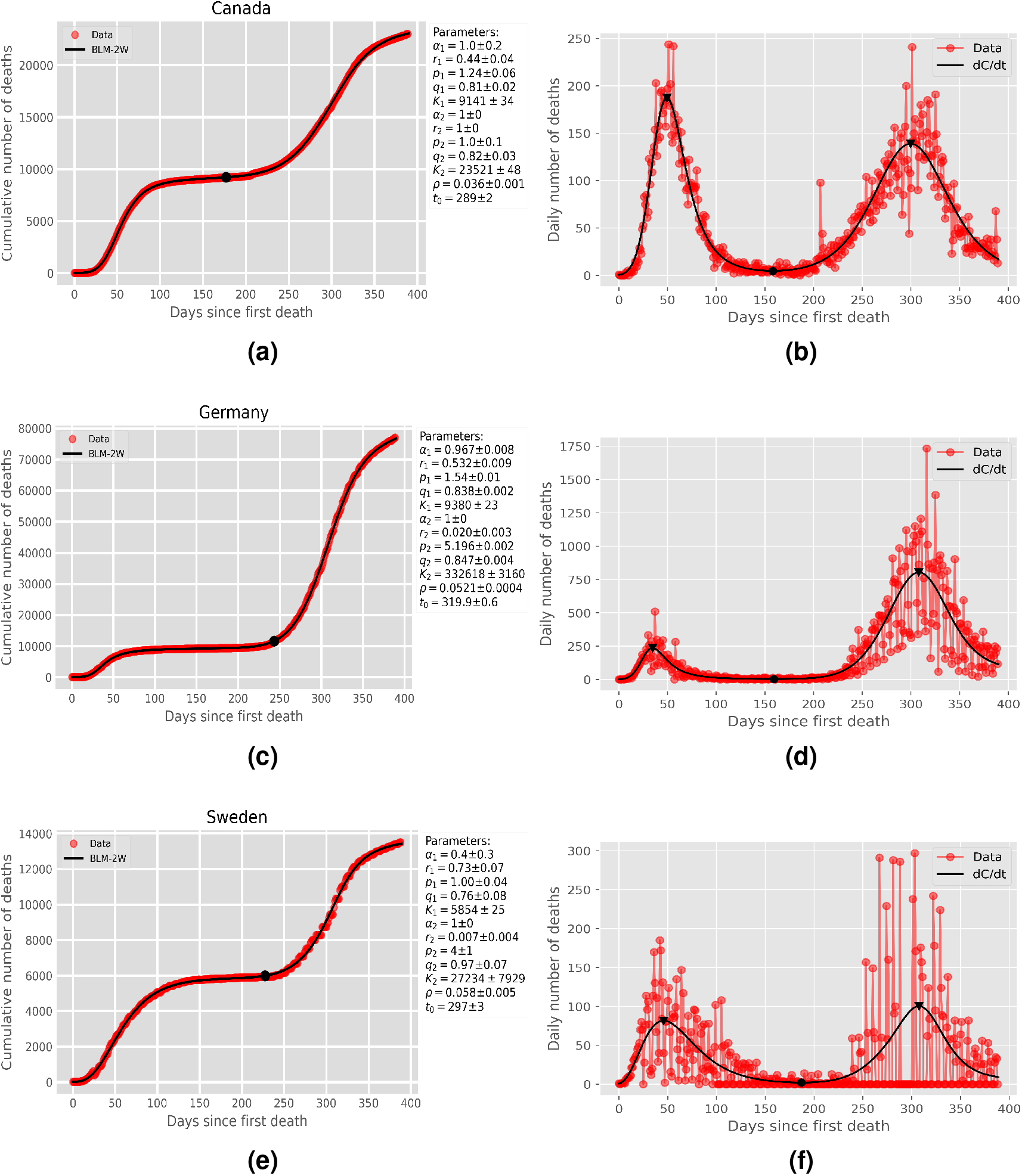
Left panels: Cumulative number of deaths (red circles) attributed to COVID-19 for (a) Canada, (c) Germany, and (e) Sweden, up to April 03, 2021. The solid curves are the best fits by the second-wave model, where the black dot in each curve represents the time, 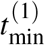, that separates the first and second waves. Right panels: Daily number of deaths for the same countries as in the corresponding left panels, where the empirical data are indicated by red circles and the solid curve represents the time derivative of the respective theoretical curve in the left panels. The maxima and minimum of the daily curves are indicated by the inverted triangles and the black dots, respectively.

In the right panels of Fig. 3 we show the daily numbers of deaths for the respective countries shown in the corresponding left panels. Here again the red circles represent the empirical data, while the black solid curves correspond to the time derivative of the theoretical curve *C*(*t*) predicted by the two-wave model, as obtained from the fits shown in the left panels of the respective figures. One sees that the theoretical daily curves are also in very good agreement with the empirical data. In particular, the model predicts remarkably well the location and general shape of both peaks (in the daily empirical curves) associated with the first and second waves, respectively. It is worth emphasizing that the numerical fits are performed only for the cumulative curves, so that the good agreement between the theoretical daily curves and the empirical daily data represents a further consistency check of the model. Note, in particular, that the two peaks in the respective daily curves in Fig. 3 are well separated by a shallow valley, corresponding to the intervening near-plateau between the two waves in the cumulative curve.

In another possible scenario, an “anomalous” second wave can develop well before the first wave has significantly subsided, causing the cumulative curve to change trend at some point in time (before it reaches a plateau) and re-accelerate again. Examples of such situations are shown in Fig. 4 for the epidemic curves of Brazil, Mexico, and South Africa, where the same symbol convention as in Fig. 3 is used. Note that in such cases there is a sort of “superposition” of two waves in the sense that a second wave-like surge appears when the daily deaths (of the first wave) are still relatively high. This implies that the two peaks in the daily curves are not well separated apart, with a relatively high valley between them, as seen in Fig. 4. Note, however, that in Brazil, contrarily to the other two countries shown in Fig. 4, the second wave has not yet reached its peak, as seen in Fig. 4b. We remark, furthermore, that among the three countries with anomalous second waves, South Africa has the least anomalous of them, in the sense that its daily curve has the lowest minimum between the two peaks; see Fig. 4f. Nevertheless, this ‘valley’ is considerably higher than those seen in the countries shown in Fig. 3, where a more standard second wave has taken place.

**Figure 4.**
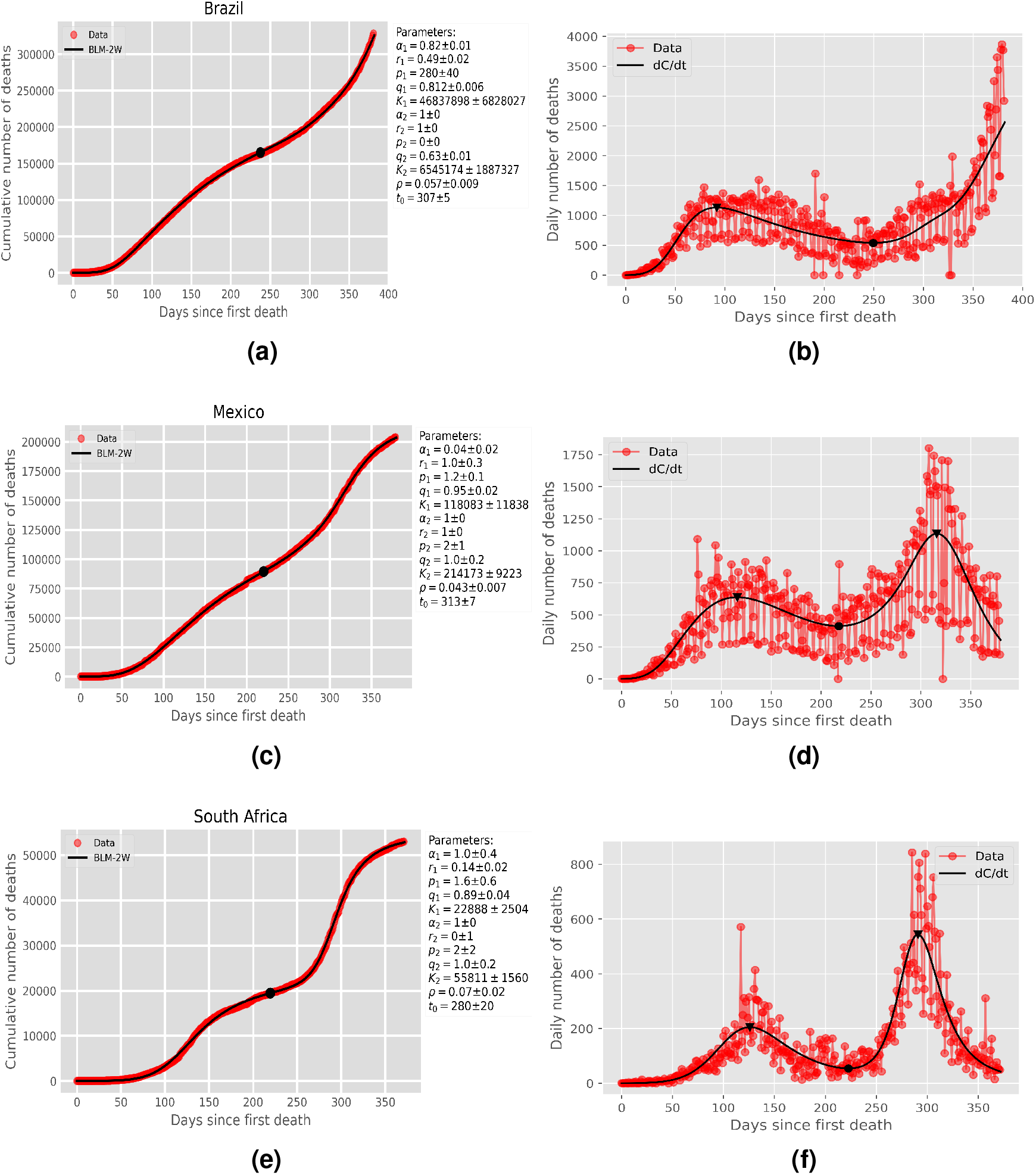
Same as in Fig. 3 for (a) Brazil, (c) Mexico, and (e) South Africa.

We have seen from Figs. 3 and 4 that our two-wave model is capable of describing very well the epidemic curves, over their entire range, for both types of second waves described above. Let us now examine countries that have three waves of infections.

### 4.2 Countries with Three Waves

In Fig. 5, we show the cumulative number of deaths for the four countries (Iran, Italy, Japan, and US) with three waves. In this figure we use the same convention as before, where the empirical data are denoted by red circles and the theoretical predictions by solid black curves. In this case the best fits are performed with the multiple-wave model given in (1) and (18), with *N* = 3. Again, one sees from the plots in Fig. 5 that the theoretical curves describe very well the empirical data for all cases shown.

**Figure 5.**
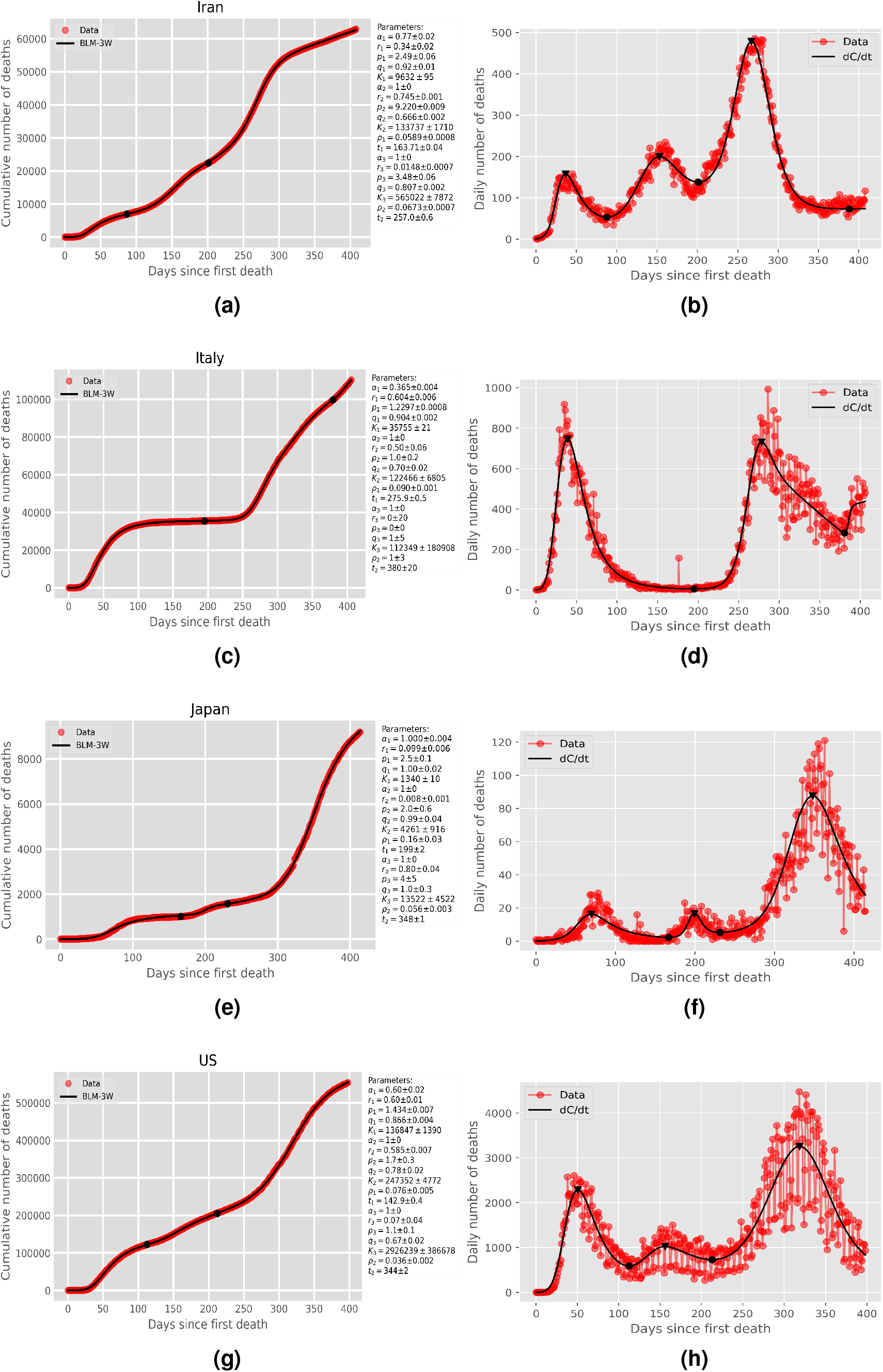
Same as in Fig. 3 for (a) Iran, (c) Italy, (e) Japan, and (g) US.

One sees from Fig. 5 that the second waves for Iran and US can be classified as being of the anomalous type (in the sense described above). In the case of Italy, the second wave has instead a more standard pattern, similar to that seen in the countries shown in Fig. 3. Japan’s epidemic curve is also of particular interest, as the second wave there started from a rather low minimum death rate (see Fig. 5f), so that on this count it resembles the standard second waves seen in other countries in Fig. 3. However, the second wave in Japan was anomalous in other ways, such as the fact that it was relatively short and overall less severe than the first one. Note, furthermore, that in all three cases of anomalous second wave among countries with three waves (i.e., Iran, Japan, and US), the third wave was the most intense one.

Another noteworthy message taken from Fig. 5 is the fact that in the cases of Iran, Japan, and US, the third waves have already clearly passed their peaks and the curves are now in a deceleration regime. The situation of Italy is, however, quite different in that the third wave is still in its early stage and the peak has not yet been reached, indicating that the epidemic curve is accelerating.

## 5 Discussion

We have seen above that the cumulative death curves of COVID-19 for many countries exhibit multiple waves of infection and death. Although a visual inspection of an epidemic curve can easily reveal whether a second or subsequent wave of infections is likely to be present, estimating more precisely when such resurgences actually begin is not an obvious task. We have argued in Sec. 3.3 that the time of occurrence of the minima in the daily curve, as predicted by our model, can be taken as an estimate for the time when the effects due to a new wave of infection start to become important. The corresponding values of such minima for each of the selected countries are indicated in Figs. 3-5 by a black dot on both the cumulative and daily curves. From inspecting these figures, one sees that the black dots do indeed represent good estimates for the separating points between successive waves.

Similarly, we can estimate the locations and intensities of the peaks of the successive waves in a given epidemic curve by computing the time of occurrence and the value of the respective maxima of the theoretical daily curve. The points of local maxima are indicated in the right panels of Figs. 3-5 by an inverted triangle. A more detailed discussion about the intensities and duration of these multiple waves is presented next.

Based on our model, we can define a measure of the intensity of a given subsequent wave, relative to the preceding one, by considering the ratio of the two successive peaks in the daily curve. More precisely, we consider the following measure for the intensity of the *k*-th wave, with *k* > 1, relative to the preceding wave:

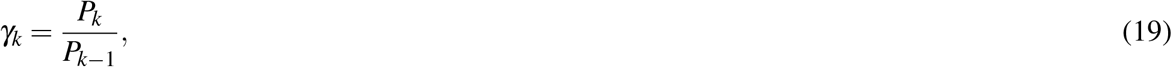

where *P*_*j*_ denotes the value of the *j*-th maximum (peak) of the theoretical daily curve. More precisely, we define

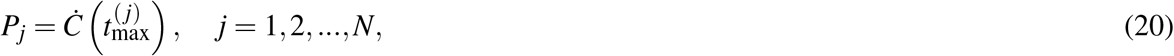

where 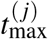 is the location of the *j*-th peak, i.e., 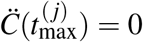 and 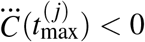, with dots denoting time derivative. For later use, we shall also define the values of the minima (heights of the valleys) of the daily curve by

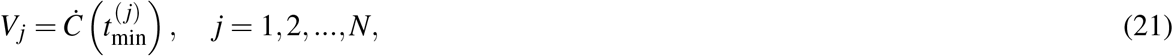

where 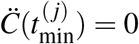 and 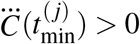.

For countries with only two waves, the values of 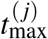 and *P*_*j*_, for *j* = 1, 2, as well of 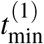, *V*_1_, and *γ*_2_ are shown in table 1. One important feature seen in this table is the fact that the minimum values *V*_1_ for Canada, Germany, and Sweden are indeed quite low (*V*_1_ ≤ 5), in comparison to what is observed for Brazil, Mexico, and South Africa, where *V*_1_ = 541, 413, 54, respectively. This confirms our findings anticipated above that the latter countries have experienced a somewhat anomalous second wave, in the sense that the daily number of deaths starts to grow again well before the first wave has been brought under control. This effect is particularly strong in Brazil (*V*_1_ = 541) and Mexico (*V*_1_ = 413), indicating that these two countries have not enforced effective measures to tame the first wave of COVID-19.

**Table 1.**
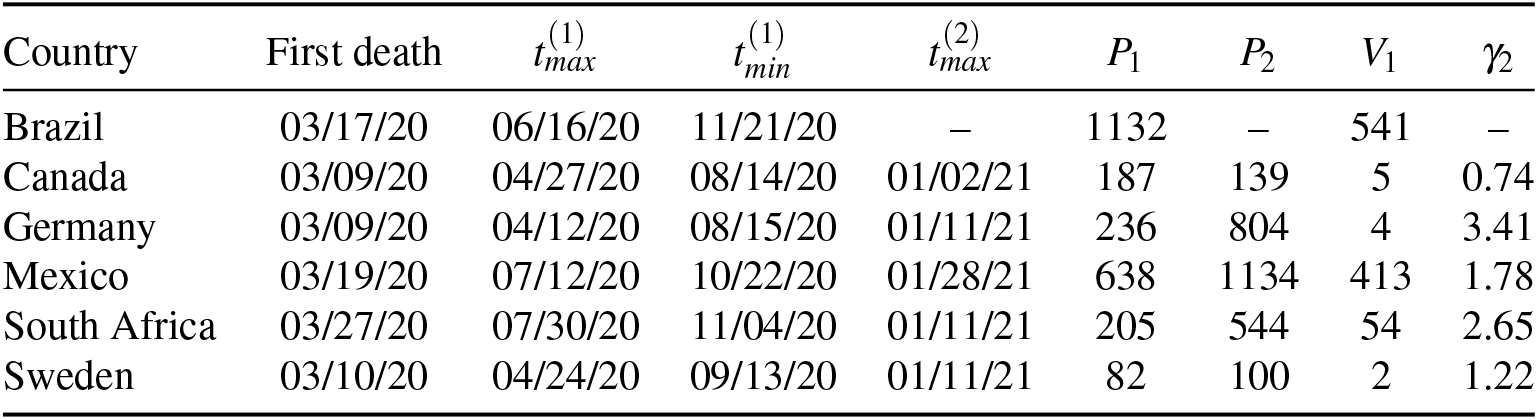
Parameters estimating the location and respective values of the maxima and minimum of the daily curves for countries with two waves.

| Country | First death | $t_{\max}^{(1)}$ | $t_{\min}^{(1)}$ | $t_{\max}^{(2)}$ | $P_1$ | $P_2$ | $V_1$ | $\gamma_2$ |
| --- | --- | --- | --- | --- | --- | --- | --- | --- |
| Brazil | 03/17/20 | 06/16/20 | 11/21/20 | – | 1132 | – | 541 | – |
| Canada | 03/09/20 | 04/27/20 | 08/14/20 | 01/02/21 | 187 | 139 | 5 | 0.74 |
| Germany | 03/09/20 | 04/12/20 | 08/15/20 | 01/11/21 | 236 | 804 | 4 | 3.41 |
| Mexico | 03/19/20 | 07/12/20 | 10/22/20 | 01/28/21 | 638 | 1134 | 413 | 1.78 |
| South Africa | 03/27/20 | 07/30/20 | 11/04/20 | 01/11/21 | 205 | 544 | 54 | 2.65 |
| Sweden | 03/10/20 | 04/24/20 | 09/13/20 | 01/11/21 | 82 | 100 | 2 | 1.22 |

Among the countries that have experienced a standard second wave, Germany had the more intense second wave relative to the first one, with a relative intensity of *γ*_2_ = 3.41; see table 1. This shows that, while Germany was quite successful in controlling the first wave of COVID-19, the relaxation of control measures during the Summer months of 2020, compounded perhaps by the lack of adherence by the population to the health authorities’ guidelines, lead to a strong resurgence of the epidemic, so much so that the great majority of deaths in Germany occurred during the second wave, as one can see from Fig. 3c.

In connection with table 1, it is also worth pointing out that among the three countries with standard second waves, the second wave started 5-6 months after the first death (namely from mid-August to mid-September, 2020). In contradistinction, for the three countries with anomalous second waves the first wave lasted longer and the second wave began only 7-8 months after the first death (namely, around the October-November, 2020). Considering that Mexico is in the Northern Hemisphere (albeit in a tropical/subtropical region), as are the countries with standard second waves in Figs. 3, the different time scales for the onset of the second waves seen in the two groups of countries in Figs. 3-4, seem to reflect the general strategies adopted in the respective countries, rather than seasonal effects. In other words, it may so be that the climate season (Summer) that coincided with the relaxation of the mitigation measures, say in Europe, played only a small part in explaining the timing of the second wave. A similar behavior was seen for the 1918 influenza pandemic, where seasonal effects were difficult to predict^38^. It might indeed be the case that the internal dynamics of the complex epidemic system, as represented by the virus propagation dynamics coupled to the population responses, inherently brings about a time scale for the occurrence of successive waves. This is an interesting possibility that deserves to be studied further.

In table 2 we show the the values of 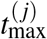 and *P*_*j*_, for *j* = 1, 2, 3, 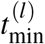 and *V*_*l*_, for *l* = 1, 2, as well as *γ*_2_ and *γ*_3_, for the countries with the three waves. Ones sees from this table that the values (*V*_1_) of the first minimum of the daily curves for Iran and US are considerably higher than those for Italy and Japan, thus confirming the ‘anomalous’ nature of the second waves in the former two countries. The second wave in the US was particularly anomalous (as was the case in Brazil and Mexico; see table 1), in the sense that the second resurgence started when the daily number of deaths was still quite high (*V*_1_ = 590 for the US). Japan’s second wave is also somewhat anomalous, as compared, say, to the countries shown in Fig. 3, but there the ‘anomaly’ has more to with the fact that the second wave had a relatively short duration of about two months, as estimated from the interval between 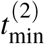 and 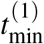, which was approximately the same duration as the second wave in the US. Furthermore, we also see from table 2 that in the three countries where the third wave has already peaked, this last wave was the most intense one, as confirmed by the fact that not only the factor *γ*_3_ but also the product *γ*_2_*γ*_3_, which gives the relative intensity of the third wave compared to the first, are all significantly greater than one.

**Table 2.** Parameters estimating the location and respective values of the maxima and minimum of the daily curves for countries with three waves.

| Country | First death | $t_{max}^{(1)}$ | $t_{min}^{(1)}$ | $t_{max}^{(2)}$ | $t_{min}^{(2)}$ | $t_{max}^{(3)}$ | $P_1$ | $P_2$ | $P_3$ | $V_1$ | $V_2$ | $\gamma_2$ | $\gamma_3$ |
| --- | --- | --- | --- | --- | --- | --- | --- | --- | --- | --- | --- | --- | --- |
| Iran | 02/19/20 | 03/26/20 | 05/16/20 | 07/21/20 | 09/07/20 | 11/11/20 | 158 | 200 | 478 | 53 | 138 | 1.27 | 2.39 |
| Italy | 02/21/20 | 03/30/20 | 09/03/20 | 11/25/20 | 03/07/21 | – | 746 | 731 | – | 5 | 281 | 0.98 | – |
| Japan | 02/13/20 | 04/22/20 | 07/28/20 | 08/30/20 | 10/01/20 | 01/25/21 | 16 | 17 | 88 | 2 | 5 | 1.06 | 5.18 |
| US | 02/29/20 | 04/19/20 | 06/20/20 | 08/03/20 | 09/28/20 | 01/12/21 | 2287 | 1020 | 3253 | 590 | 730 | 0.45 | 3.19 |

We have seen above that, among the ten countries selected here, six of them (Brazil, Iran, Japan, Mexico, South Africa, and US) have developed anomalous second waves. It is reasonable to argue that such an anomalous behavior is most likely an indication of the fact that mitigation measures were relaxed prematurely (i.e., before the first wave was brought under control). In contrast, in countries that experienced a standard second wave (even if eventually a third wave ensued, as in the case of Italy), the second resurgence started only after a relatively long period during which the epidemic was kept under control, as indicated by a low daily death rate over this period; see, e.g., the long and shallow valleys between the first two peaks in Figs. 3b, 3d, and 5d.

One important and worrisome aspect of our results above is the fact that, regardless of the different evolution patterns in the countries considered here, in all cases the subsequent waves (independently of their number) accounted for the majority of the mortality toll. This is a clear and sad reminder that the local populations and health authorities must remain vigilant at all times, from beginning to the end of the pandemic, as the risk of a resurgence is always present for as long as the epidemic is not brought under effective control worldwide. In fact, there are already some countries that are undergoing a fourth resurgence of the epidemic. An example, for illustrative purposes only, is shown in Fig. 6 for Serbia, where one clearly sees four distinct waves of infections of increasing intensity. Our *N*-wave model, with *N* = 4, could in principle be straightforwardly applied to such a curve, but in this case the numerical task of fitting the parameters becomes more challenging, due to the large number of parameters to be determined (26 of them for *N* = 4). Hence in the present study we have restricted our analysis to COVID-19 mortality curves that display at most three waves.

**Figure 6.**
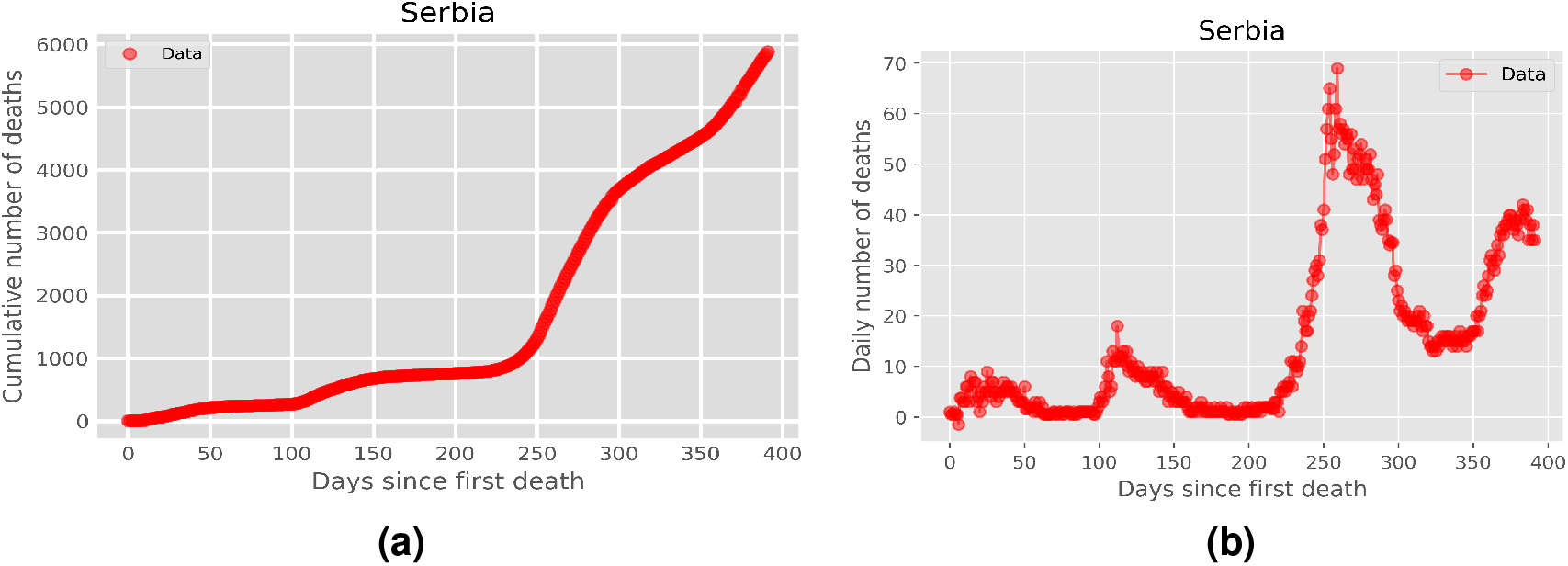
Cumulative (left panel) and daily (right panel) number of deaths attributed to COVID-19 for Serbia up to April 15, 2021, where one can clearly identify four distinct waves of infection.

## 6 Conclusion

In this paper, we have studied the dynamics of the second and third waves of infections by the novel coronavirus. To this end, we have introduced a generalized logistic model with time-dependent parameters to analyze the COVID-19 fatality curves of ten countries from five continents. Not only the theoretical curves are in excellent agreement with the empirical data for all cases considered, but they also allow us to infer predictions about the location and severity of the first and subsequent waves. For instance, by estimating the starting point (in terms of the minimum daily death rate) and duration of a given subsequent wave, we have argued that they can be qualitatively classified as being either of a standard type (i.e., one that follows after the previous wave has nearly subsided) or of an anomalous nature (i.e., where the resurgence of infections takes places while the preceding wave was still developing).

Furthermore, we have argued that the occurrence of such anomalous waves is the consequence of a premature relaxation of the control measures by government and health officials. Similarly, in countries that have experienced a standard second wave, a sort of “paradox of success”^39^ seem to have been at play, whereby early success in controlling the first wave of COVID-19 might have led to a false impression that “the worst was behind,” thus stimulating a relaxation of voluntary or enforced measures beyond what would be desirable. In the present study we have analyzed only ten representative countries, as our main objective was to introduce and validate our multiple-wave model. Nevertheless, we expect that the trends identified here should hold in general.

The results reported in the present paper are relevant both from a mathematical viewpoint, in that they show that additional care is needed when modelling multiple-wave epidemics, and from a practical perspective, for they may help policymakers and health authorities in devising strategies to battle the disease during all of its waves. Here we have restricted our analysis to second- and third-wave effects, but our model can in principle be applied to any number of multiple waves.

## Data Availability

The data used in this study were obtained from the data base made publicly available by the Johns Hopkins University

